# Development of a universal single-item therapeutic empathy scale and validation of the patient-reported version

**DOI:** 10.64898/2026.03.28.26349594

**Authors:** Amber Bennett-Weston, John Maltby, Kamlesh Khunti, Char Leung, Daya Narwal, Patience Otoo, Benita Iyadi-Wilson, Jeremy Howick

## Abstract

**Background:** Therapeutic empathy improves patient and practitioner outcomes, yet existing measures are often lengthy, conceptually inconsistent, and cannot be easily compared across respondent groups. Brief, universal measures (usable by patients, practitioners, students, and observers) are lacking. We therefore developed a universal single-item scale and conducted psychometric testing of the patient-reported version.

**Methods:** Following best-practice, we used a three-phase approach: (1) item development; (2) pre-testing the scale by obtaining expert panel feedback (n=9) and conducting cognitive interviews with stakeholders (n=35); and (3) scale validation in an international patient sample (n=521) assessing convergent, discriminant, and known-groups validity. Validation involved assessing correlations with the Consultation and Relational Empathy (CARE) measure and clinical neutrality measure, and by assessing differences in scores by patient ethnicity.

**Results:** We developed two versions (pictorial and text-based) of each scale. Expert feedback and cognitive interviews confirmed content and face validity. Pictorial and text-based versions showed high convergent validity with the CARE measure (r=0.761 and r=0.838, both p<0.001), and discriminant validity with a clinical neutrality measure (r=0.131 and r=0.139, p=0.003 and p=0.001, respectively). Correlations with the CARE measure remained high (r>0.70) and statistically significant (p<0.001) across patient gender, ethnicity, and practitioner type. Ethnic minority patients rated practitioner empathy lower than White patients (pictorial p=0.057; text-based p=0.033), demonstrating known-groups validity. Patients rated doctors’ empathy higher than other healthcare practitioners’ (p=0.001 for both pictorial and text-based); there were no significant differences in empathy scores by patient gender.

**Conclusions:** We developed the first universal single-item therapeutic empathy measure and demonstrated validity for the patient-reported versions. The scale is brief, accessible, and applicable to clinical practice, education, and research. Further research should validate practitioner-, student-, and observer-reported versions, and assess predictive and cross-cultural validity. This robust tool can support patient-reported routine measurement of therapeutic empathy and contribute to improving patient and practitioner outcomes.

## 1. Introduction

Therapeutic empathy (sometimes called “clinical empathy”) is a cornerstone of healthcare practice, research, and education [1-3]. For patients, therapeutic empathy can reduce pain, depression, and anxiety while improving safety and satisfaction with care [4-6]. For practitioners, therapeutic empathy is associated with reduced burnout and greater job satisfaction [7, 8].

Challenges in defining therapeutic empathy have, until recently, inhibited research in this area. However, a recent systematic review and thematic analysis of definitions identified six common components across definitions: exploring and understanding the patient’s perspective, reaching shared understanding, feeling, taking therapeutic action, and maintaining boundaries [9-11].

Despite conceptual advances, approaches to measuring therapeutic empathy remain highly variable [12]. Systematic reviews [13-15] have identified multiple practitioner self-reported, patient-reported, and observer-reported measures [16-23], although observer measures are rarely used because of reliability concerns [13]. Aside from concerns about the reliability and validity of existing measures, one of their main limitations is that none were designed for use across different respondent groups [13-15]. For example, the Jefferson Scale of Empathy (JSE) was initially developed and validated for use by physicians [24], while the Consultation and Relational Empathy (CARE) measure was designed specifically for patients to assess their practitioner’s empathy [25].

Different therapeutic empathy scales also appear to measure a variety of incompatible constructs [12-15]. For example, although the JSE defines empathy as involving cognitive understanding and therapeutic action [22], the measure focuses largely on cognitive elements [15]. Meanwhile, the CARE measure captures the relational aspects of empathic patient-practitioner interactions (for example, how well the practitioner listened), with less consideration of cognitive components of therapeutic empathy. Relatedly, observer-reported measures such as the Therapist Empathy Scale (TES) focus on the emotional aspects of empathy [13].

This variation in measures matters. If we wish to test whether an empathy intervention is effective for improving therapeutic empathy (and subsequently, patient and practitioner outcomes), we need to know which measure is optimal. Relatedly, if we wish to measure whether our training interventions [26] are effective at teaching therapeutic empathy to medical students, then it is important for observer-rated empathy measures to be suitably validated [13]. Moreover, the lack of conceptual coherence between measures makes it difficult to compare therapeutic empathy scores across perspectives or determine whether different scales measure the same construct [27], potentially contributing to discrepancies between practitioner self-ratings and patient ratings [28-30].

In an attempt to address this problem, some existing scales have undergone post-hoc adaptation for other groups. For example, the JSE has been adapted for use by patients [31], and the CARE measure has been adapted for use by practitioners [32]. However, such post-hoc adaptations repurpose instruments not specifically designed for these groups, risking misalignment between item content and the construct as experienced by different respondents. This, in turn, can compromise content validity and conceptual equivalence [33, 34].

Another problem with existing scales is their complexity. Many have over 15 items and include negatively worded questions [16-24]. This can increase survey fatigue and risks incomplete responses [35, 36]. Even the widely used 10-item Consultation and Relational Empathy (CARE) measure [25] may be burdensome in some time-pressured clinical settings [37].

Single-item scales reduce time burden while offering effective measurement, and higher response and completion rates [38-44]. They are particularly suited for time-constrained clinical settings where routine measurement is required [45, 46]. Single-item scales also reduce the risk of criterion contamination [40], improve face validity by removing ambiguous items [40, 47, 48], and simplify translation across diverse populations [49, 50]. While concerns have been raised about the reliability and sensitivity of single-item scales, they have not been substantiated [39, 51]. Although several single-item therapeutic empathy scales have been proposed [52-55], none have been validated.

It is therefore important to have a single-item, validated therapeutic empathy scale that measures the same construct and that is designed from the outset for use by patients, practitioners, students, and observers [13-15]. We describe this as a “universal” scale. Developing such a scale is especially important given recent research showing the importance of empathy for enhancing patient satisfaction [4], safety [5], and practitioner wellbeing [7].

### 1.1. Objectives

This study has two objectives:

1. To develop the first “universal” single-item measure that integrates all six components of therapeutic empathy [11] and is suitable for use by patients, practitioners, students, and observers.
2. To test the construct validity of the patient-reported version.

## 2. Materials and Methods

### 2.1. Study design

We conducted a mixed-methods study to develop a universal single-item scale of therapeutic empathy, and to psychometrically test the patient-reported version. Following established best-practice for developing and testing scales in health, social, and behavioural research [33, 34, 56], we followed three phases: 1) item development, 2) scale development, and 3) scale evaluation (Figure 1) [33]. We reported the study following recommendations for reporting scale development and testing [56].

**Fig 1.**
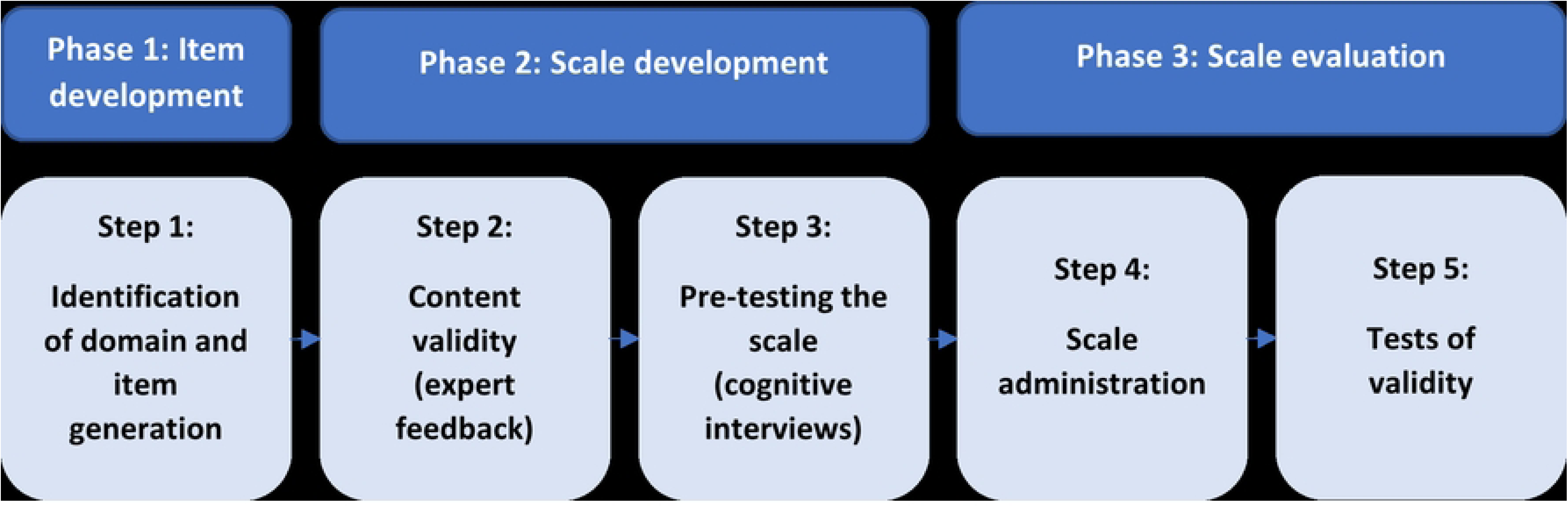
Study flow diagram.

### 2.2. Sampling and recruitment

We recruited three groups of participants:

1. Stakeholders with expertise on therapeutic empathy in healthcare research, practice or education to assess content validity of the universal scale.
2. Stakeholders from each target population (patients, practitioners, observers) to assess face validity and pre-test the universal scale.
3. Patients to complete the final patient-reported scale to enable tests of validity.

Sample sizes for each participant group were chosen to meet established standards [33, 34, 56]. All participating patients were offered remuneration in line with national guidance [57].

#### 2.2.1. Stakeholders with expertise on therapeutic empathy to assess content validity

We aimed to recruit a minimum of five expert panellists (including patient representatives, therapeutic empathy researchers and educators, and healthcare practitioners) to assess content validity [33, 34]. We purposefully sampled [58] participants with relevant expertise in across medicine, nursing, patient advocacy, medical ethics, and scale development, of different ages, genders, and ethnicities, and from different continents [34, 59], and invited them to participate via email between 2^nd^ June 2025 and 2^nd^ July 2025. All participants provided written informed consent to participate. We defined expertise as having made a substantial contribution to the field of therapeutic empathy either through research, teaching, or practice [60].

#### 2.2.2. Stakeholders from each target population to assess face validity and pre-test the scale

We aimed to recruit at least 20 patients, practitioners, healthcare students, and observers (healthcare educators, carers) to participate in cognitive interviews to pre-test all versions of the universal scale. The final sample size was determined by reaching meaning saturation: the point at which we had a rich understanding of participants’ views and experiences of using the scale [61]. We purposefully selected patients of varied ages, ethnicities, genders, and healthcare experiences across the UK via local patient groups and the NIHR People in Research platform [62]. We also recruited a diverse sample of practitioners, students, and educators from multiple professions in the East Midlands, via a Medical School and a Healthcare School, and their associated NHS Trusts. Recruitment took place between 15^th^ July 2025 and 30^th^ July 2025. All participants provided written informed consent to participate.

#### 2.2.3. Patients to complete the final patient-reported scale to test validity

We aimed to recruit at least 200 patients aged ≥18 years, of any gender or ethnicity, who had seen a healthcare practitioner in any setting within the past year to complete the final patient-reported version of the universal scale. We recruited an international convenience sample of patients using online survey recruitment platform Prolific [63]. We supplemented this by recruiting in-person from General Practices, Maternity Services, Infectious Disease, and Geriatrics clinics across an NHS Trust and Primary Care Network (PCN) in the East Midlands of the UK to capture patients experiencing digital exclusion or limited digital literacy. Clinics were selected for their diverse populations [64, 65]: the General Practices served the most deprived ward within their PCN [66]; the Maternity Services served women and families in the UK’s first plural city; the Geriatric Clinic served older people; and the Infectious Disease Clinic served many homeless people. A screening question based on previous research [67] assessed digital exclusion and illiteracy (S1 Appendix), and at least ten percent of the sample was recruited in-person to reflect the estimated proportion of digitally excluded or illiterate patients in the UK [68, 69]. Recruitment took place between 1^st^ September 2025 and 15^th^ November 2025. All participants provided written informed consent to participate.

### 2.3. Procedures

#### 2.3.1. Identification of domain and item generation

Therapeutic empathy was the construct of interest, and recent systematic reviews indicated that existing measures did not meet the aims of the scale we intended to develop; none were designed for universal use by patients, practitioners, students, and observers, and many lacked user-friendliness [13-15]. We therefore started scale development, using previous qualitative research on patients’ and practitioners’ lived experiences of therapeutic empathy [70-72] and a recent systematic review which identified six common components of the concept (S2 Appendix) as our foundation for item generation [10, 11].

To support the accessibility of the scale across different respondent groups with varying literacy levels [72], we adopted several strategies: first, we used Plain English language and aimed for a reading age of below 13 years [73]. To achieve this, we assessed the initial scales using readability tests: the Flesch-Kincaid Grade Level and Flesch Reading Ease Test [74]. Second, following previous studies [25, 75], we aimed to develop two versions of the scale for each target population: a pictorial version and a text-based version. Third, we opted for unipolar five-point measures to reduce conceptual ambiguity [33, 76].

#### 2.3.2. Content validity

To assess content validity [33], our expert panel reviewed the initial versions of the universal scale, along with the study definition of therapeutic empathy [11], and provided anonymised feedback on clarity and relevance via an online survey (S3 Appendix). This feedback informed scale refinement.

#### 2.3.3. Pre-testing the scale

We assessed face validity and pre-tested the scale through cognitive interviews [33] to explore how potential respondents interpreted and formulated answers to the scale, steered by a semi-structured topic guide (S4 Appendix) [77]. We offered participants the choice of conducting their interview either online (via Microsoft Teams version 25306.804 [78]) or in-person (at the host institution). All interviews were conducted by at least one researcher (from either ABW or DN), and were audio-recorded and transcribed. Using a light-touch approach to thematic analysis [79], we summarised the interviews and used this to further refine each scale. We conducted the cognitive interviews in rounds of at least five participants (including a mix of participants from each respondent group) to enable us to modify and test each version of the scale iteratively [33, 77, 80], and continued until we reached meaning saturation [61]. By this point we had made exhaustive modifications, leaving us with the final versions of the scale.

#### 2.3.4. Survey administration

We administered the final versions of the patient-reported scale to a larger sample of patients online via Prolific [63] and in-person at our NHS sites. We first tested the patient-reported version as this has been identified as a priority [72]. To measure convergent and divergent validity (see below), we administered the CARE measure [25] and a question about clinical neutrality (S5 Appendix), which we adapted from previous research [24].

#### 2.3.5. Tests of validity

Typically, establishing internal reliability and examining latent structure (for example via internal consistency indices and exploratory factor analysis) would be the next step in scale evaluation [33]. However, these approaches are not applicable to a single-item measure [33, 41, 51, 81]. Moreover, test-retest reliability is primarily used to assess the stability of an underlying trait. This approach is unlikely to be meaningful because therapeutic empathy is inherently interaction- and context-dependent, rather than a stable characteristic of a single consultation [82, 83].

Therefore, in the final step we assessed construct validity, including convergent validity, discriminant validity, and known-groups validity [84] using R Version 4.5. We assessed convergent validity by measuring correlations between the two versions of our scale and one other well-established patient-reported measure of therapeutic empathy: the CARE measure [22, 25]. A high correlation would represent high convergent validity. To further explore the robustness of any correlations between our scales and the CARE measure [25], we repeated the correlational analyses across key demographic subgroups, including patient gender, ethnicity, and type of practitioner they were treated by [33].

We assessed discriminant validity by measuring correlations between our scale and a measure of a theoretically unrelated construct: clinical neutrality. This concept has been used in previous therapeutic empathy scale validation studies to assess discriminant validity [24]. A low correlation would indicate strong discriminant validity, by demonstrating that our measure does not correlate too highly with an unrelated construct [33].

We used Spearman’s Rank correlation coefficient to assess convergent and discriminant validity given the ordinal nature of the data, and interpreted correlation strength as follows: (-) 0.90 to 1.00 indicates a very high correlation, (-) 0.70 to 0.90 indicates a high correlation, (-) 0.50 to 0.70 indicates a moderate correlation, (-) 0.30 to 0.50 indicates a low correlation, and below (-) 0.30 indicates a negligible correlation [85]. In the context of convergent validity specifically, although a correlation coefficient of (-) 0.50 is broadly considered “sufficient”, it is recommended that a correlation coefficient of (-) 0.70 should be used as a benchmark to indicate that two scales genuinely measure the same construct [86].

Known-groups validity (the extent to which a scale can discriminate between two or more groups theoretically expected to differ on the variable of interest) was assessed by comparing scale scores across groups expected to differ in their therapeutic empathy ratings using the Kruskal-Wallis test [33]. Research shows that perceptions and experiences of empathy in healthcare are influenced by ethnicity [59, 87] and that specifically, patients from ethnic minority groups rate practitioner empathy lower than those from non-minority ethnic groups [88]. Accordingly, we conducted tests for known-groups validity between patients of different ethnicities.

To supplement our tests of validity, and following previous research [25], we conducted exploratory demographic comparisons between empathy ratings by patient gender and the type of practitioner they were treated by.

### 2.4. Ethical considerations

Ethical approval for this study was received on 28/05/2025 from the Health Research Authority (IRAS project ID: 350113; REC reference: 25/WS/s0067). All participants provided written informed consent to participate.

## 3. Results

### 3.1. Identification of domain and item generation

Having identified our domain – therapeutic empathy – we generated a single item scale, taking inspiration from the findings of previous research describing the nature of the construct [11, 70-72]. For each respondent group, we generated two versions of the scale: a pictorial measure using smiley-faces (ranging from not at all happy to very happy) as the response options, similar to the visual analogue scales used to measure pain [89, 90], and a text-based measure ranging from “not at all empathic” to “extremely empathic” (see Figure 2 for initial patient-reported versions and S6 Appendix for other versions).

**Fig 2.**
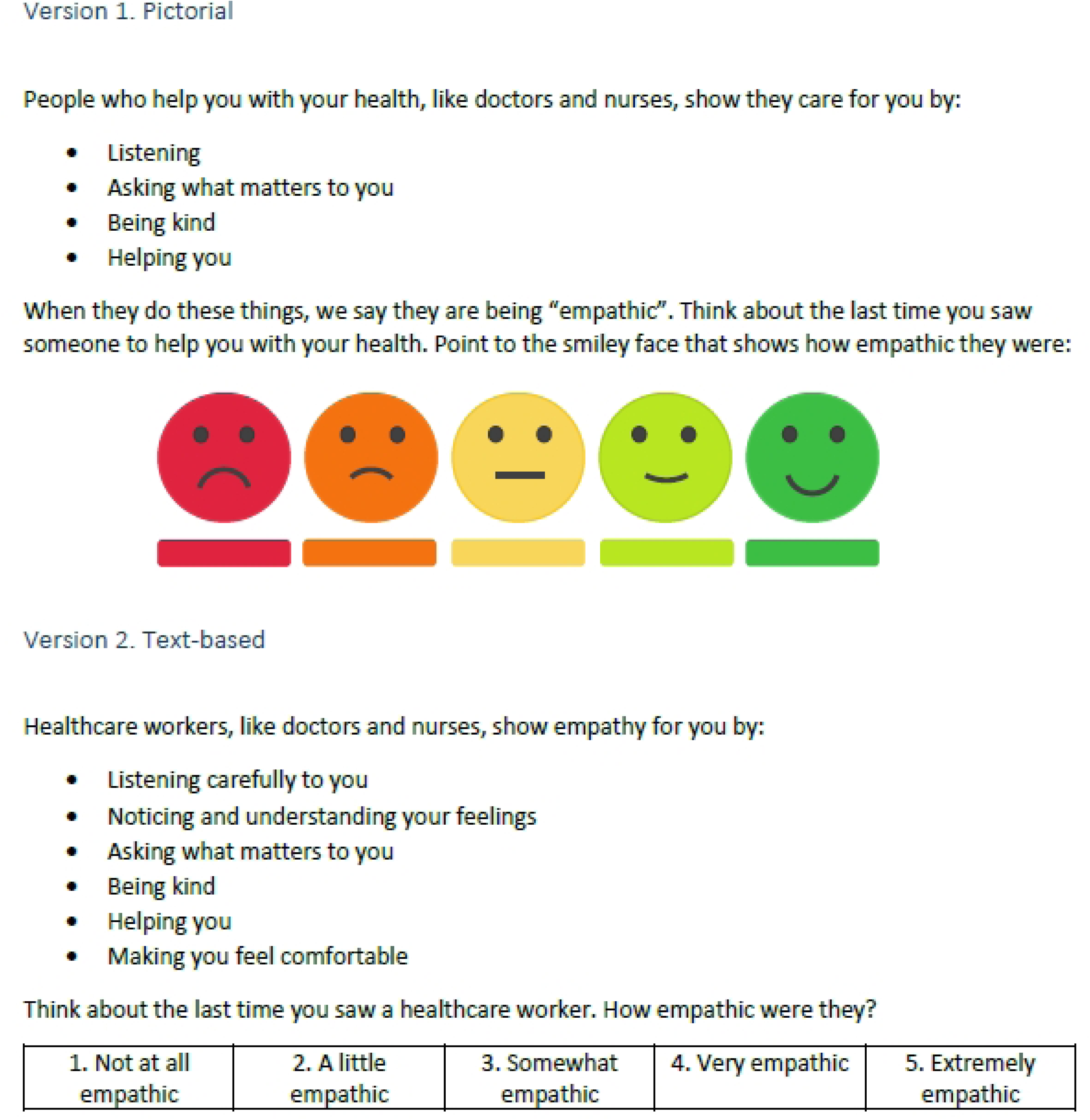
Initial patient-reported versions of the universal single-item therapeutic empathy scale.

Flesch-Kincaid Grade Level scores (range 2.8 - 7.5) and Flesch Reading Ease Test scores (range 49.0 - 88.8) confirmed that the readability of the initial versions of the scale were suitable for the intended reading age of below for 13 years for each respondent group (S7 Appendix).

### 3.2. Content validity

In July 2025, nine participants joined our expert panel to assess the content validity of the initial versions of the scale (67% female, 78% white, n=2 patient representatives, n=9 healthcare practitioners and educators, see S8 Appendix). Their feedback was positive; they described the scales as being easy to understand by each respondent group. Many expert panel members also believed that the description of therapeutic empathy provided was relevant to everyday clinical practice. They made suggestions for improving the scale, including refining the wording to better align with our definition of empathy and removing references to any single type of healthcare practitioner (for example, doctors or nurses), which we used to make modifications (see S9 Appendix for a detailed description of participant feedback and modifications made at each stage of development). The modified versions were pre-tested in cognitive interviews.

### 3.3. Pre-testing the scale

In July and August 2025, 35 participants (comprised of patients, healthcare practitioners, healthcare students, and observers such as healthcare educators and carers), 49% female, 69% white, see S10 Appendix) participated in cognitive interviews lasting between 30 and 40 minutes. Three rounds of interviews were conducted; 15 participants took part in round one, 10 participants in round two, and 10 participants in round three. Each participant took part in one interview; 11 interviews were conducted in-person, 25 took place online. Several participants had dual roles (for example, as patients and carers, or as healthcare practitioners and educators), and were able to pre-test more than one version of the scale.

In all three rounds of cognitive interviews, most participants endorsed the single-item nature of the scales, explaining that they were easy to understand and quick to complete, often taking less than two minutes. Most participants (including healthcare practitioners and students) preferred the pictorial version of the scale, finding it more engaging and even faster to complete than the text-based version. Minor modifications to wording were suggested (S9 Appendix). After the third round of interviews, we had reached meaning saturation [61] and stopped interviewing. We modified the scales once more, leaving us with the final versions ready to be tested for validity (see Figure 3 for final patient-reported versions and S11 Appendix for other versions).

**Fig 3.**
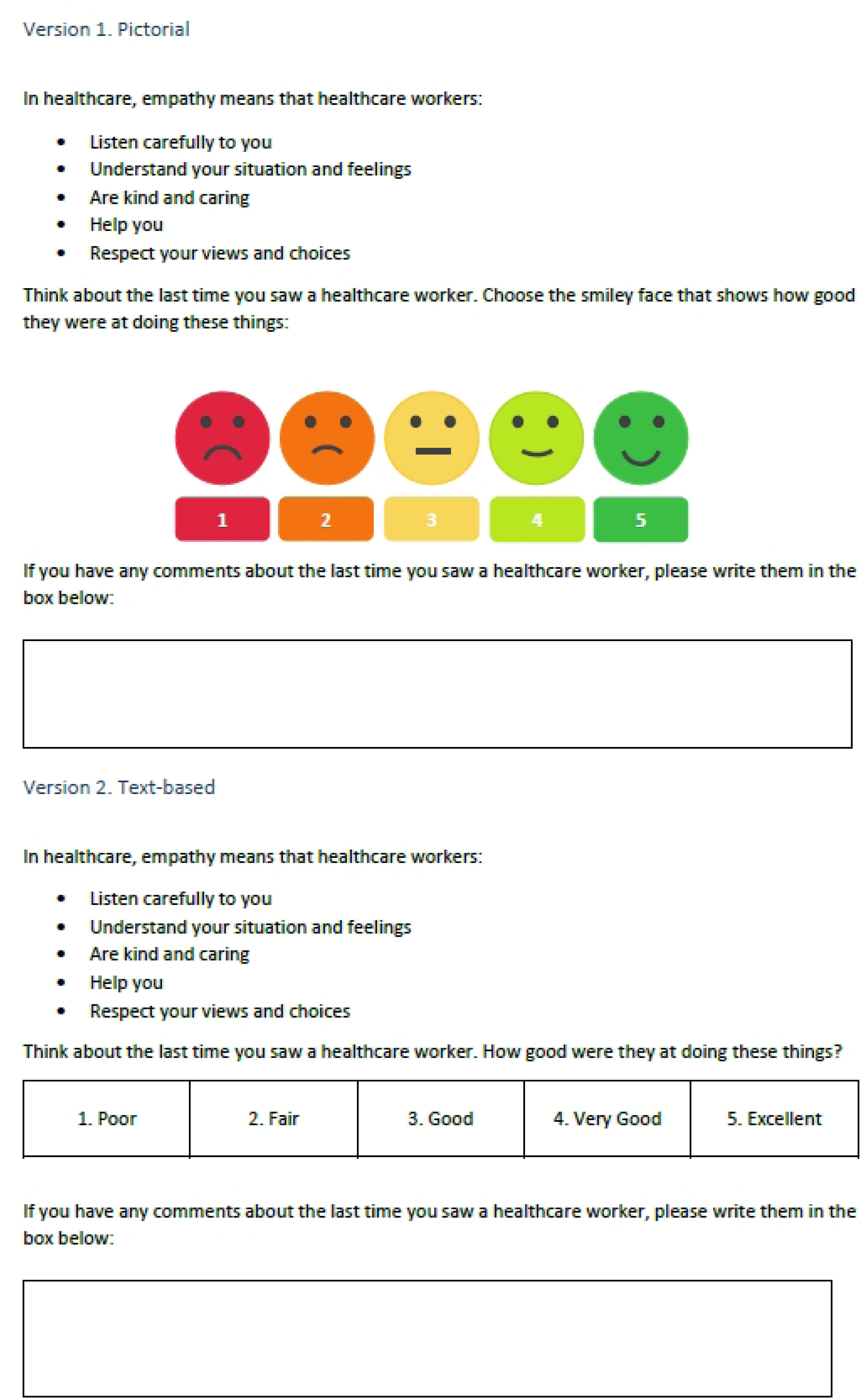
Final patient-reported versions of the universal single-item therapeutic empathy scale.

### 3.4. Scale administration and test of validity

Between September and November 2025, 521 patients completed our newly developed scales, along with the CARE measure [25] and a measure of clinical neutrality [24]. Participants were aged 18–84 years (M = 40.6, SD = 15.3). Of the sample, 274 (54%) identified as female, 227 (45%) as male, and three (1%) as other. Most participants identified as White (n = 333, 64%), followed by Black, Black British, Caribbean, or African (n = 132, 25%), Asian or Asian British (n = 27, 5%), mixed or multiple ethnic groups (n = 20, 4%), and other ethnicities (n = 7, 1%). Participants were from Europe (n = 369, 71%), Africa (n = 115, 22%), North America (n = 17, 3%), Asia (n = 15, 3%), Australia (n = 4, 1%), and South America (n = 1, <1%). Most respondents rated therapeutic empathy in primary care (n = 350, 67%), with the remainder referring to secondary (n = 114, 22%) or tertiary care (n = 57, 11%). In total, 394 participants (76%) rated a medical doctor and 127 (24%) rated an allied health professional. Finally, 109 participants (21%) were recruited in person from participating NHS Trusts, with the remainder recruited online.

High convergent validity with the CARE measure was achieved for the pictorial (r = 0.761, p < 0.001) and text-based scales (r = 0.838, p < 0.001). Strong convergent validity was also observed between the pictorial and text-based scales and the CARE measure across subgroups, including females (pictorial: r = 0.776, p <0.0001; text-based: r = 0.846, p <0.0001) and males (pictorial: r = 0.751, p <0.0001; text-based: r = 0.832, p <0.0001), White patients (pictorial: r = 0.787, p <0.0001; text-based: r = 0.873, p <0.0001) and non-White patients (pictorial: r = 0.703, p <0.0001; text-based: r = 0.748, p <0.0001), and patients treated by doctors (pictorial: r = 0.750, p <0.0001; text-based: r = 0.843, p <0.0001) and patients treated by other healthcare practitioners (pictorial: r = 0.770, p <0.0001; text-based: r = 0.797, p <0.0001).

Similarly, both the pictorial (r = 0.131, p = 0.003) and text-based (r = 0.139, p = 0.001) versions demonstrated strong discriminant validity through negligible correlations with a measure of clinical neutrality [24]. We also found evidence of known-groups validity based on patient ethnicity [33]; we observed differences in median therapeutic empathy ratings between patients from non-minority ethnic groups and patients from ethnic minority groups. Patients of White ethnicity assigned higher therapeutic empathy ratings to their practitioners than patients from other ethnic groups. Although statistical significance differed slightly between the pictorial and text-based versions (p = 0.057 and p = 0.033, respectively), both showed the same directional pattern. Figure 4 summarises the results of our statistical tests of validity.

**Fig 4.**
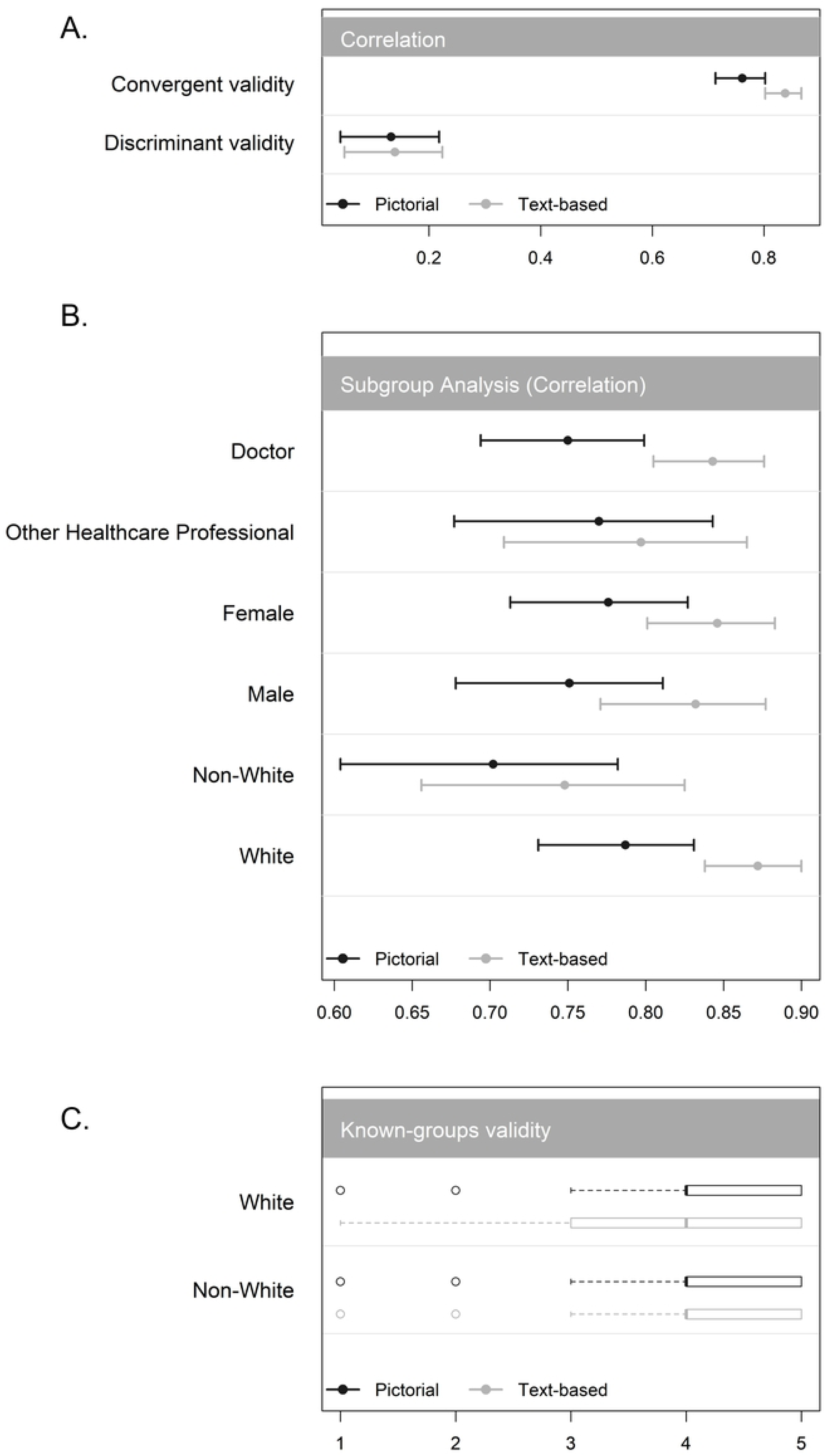
Results of statistical tests of validity.

Our exploratory demographic comparisons showed statistically significant differences between patients’ ratings of doctors’ empathy versus other healthcare professionals’ empathy for the pictorial (p = 0.001) and text-based (p = 0.001) versions of the scale, with ratings of doctors’ therapeutic empathy higher in both cases. We found no statistically significant difference in patients’ ratings of therapeutic empathy by gender on the pictorial (p = 0.625) and text-based (p = 0.698) scales.

## 4. Discussion

### 4.1. Summary of findings

We developed and validated the first universal single-item measure of therapeutic empathy. Both pictorial and text-based patient-reported versions demonstrated strong convergent validity with the CARE measure, negligible correlations with an unrelated construct, and sensitivity to known group differences. These findings suggest that a carefully designed single-item measure can provide a brief yet robust assessment of therapeutic empathy suitable for routine use in healthcare settings.

### 4.2. Comparison with other evidence

Patients from minority ethnic groups rated practitioner therapeutic empathy lower than White patients, consistent with the findings of previous research [88] and broader evidence of inequities in empathic healthcare [87]. Although statistical significance differed between the pictorial and text-based versions, the consistent direction of effects across both versions of the scale supports known-groups validity [33].

Also aligning with previous research [88], we did not find any differences in therapeutic empathy ratings between patients of different genders. This finding supports the gender invariance of the measure and indicates that it is unlikely to be biased by gender-related expectations. While previous research [91] shows that patients assign lower therapeutic empathy scores to doctors than other healthcare practitioners, in our study doctors were rated more highly than other practitioners for therapeutic empathy. It is possible that this finding reflects a sampling imbalance, as doctors were over-represented relative to other healthcare professionals. Equally, it is possible that this demographic difference has modified over recent years, given the growing recognition of the value of therapeutic empathy in healthcare [4, 5, 7].

Many existing therapeutic empathy measures overemphasise particular components of the concept or diverge from their underpinning definitions, like the JSE [22] which emphasises cognitive aspects of therapeutic empathy at the expense of affective or action-based aspects [15, 22]. In contrast, our scale captures the full breadth of therapeutic empathy through its grounding in the concepts core components [10, 11]. Combining these components into a single descriptive item may offer several psychometric advantages over multi-item scales, including reduced criterion contamination, higher face validity, superior completion rates, lower response burden, while demonstrating convergent validity with multi-item measures [40-44, 47]. Indeed, the high convergent validity of both pictorial and text-based scales with the CARE measure [25] in our study reinforces the growing consensus that one well-crafted item can be both psychometrically sound and conceptually coherent [41-44]. These arguments are strengthened by research showing that patients’ overall perceptions of empathic care better predict positive outcomes than the relative weighting of its individual components [92, 93]. Finally, our finding that a simple scale could demonstrate validity and convergence with more complex measures also dovetails with Gigerenzer’s findings that “fast and frugal” methods can be as effective (or more effective) than complex instruments [48].

### 4.3. Strengths and limitations

Key strengths of this study include adherence to best-practice methodological guidance and sustained stakeholder involvement to support content and face validity [33]. Moreover, psychometric testing in a large international and demographically diverse patient sample offers broader generalisability than many previous empathy scale validation studies which are often limited to patients from single healthcare organisations [25, 94]. Indeed, established guidance suggests that such results sufficiently meet, and even exceed, the criteria for validating a new measurement tool, with evidence of content, face, and various forms of construct validity [33, 95].

Our study also has limitations. It was not possible to assess the test-retest reliability of our scale due to the dynamic and context-dependent nature of therapeutic empathy [33, 41, 51, 81]. Relatedly, validation was limited to the patient-reported version; further testing is required to confirm the “universal” nature of the scale in practitioners, students, and observers. Finally, despite our international patient sample, Europe and Africa were over-represented relative to other continents, potentially limiting our ability to capture the cultural nuances influencing empathy ratings.

### 4.4. Implications for research and practice

Given its brevity and validity, this scale has strong potential for routine use in clinical, educational, and research settings. In practice, the scale offers a rapid, low-burden method for routine monitoring of patient experience and subsequently informing service-level quality improvement. In education, including continuing professional development, the scale could support formative assessment of, and reflection on, communication skills. This could be achieved by averaging several ratings on our scale, similar to methods deployed when using the CARE measure for appraisal or revalidation [25].

In addition to validating practitioner-, student-, and observer-reported versions of the scale in diverse healthcare contexts, further research is needed to continue testing the patient-reported version in different samples, languages, and countries. Additionally, future research can use this scale for predictive validity to check whether scores measured on this scale are associated with, or can predict, patient and staff outcomes. Finally, this study conceptualises therapeutic empathy as an attribute of the practitioner, as reflected in patient-reported evaluations. However, it is possible that patients also influence how therapeutic empathy is expressed and perceived in clinical encounters. Further research is therefore warranted to examine the role of the patient in shaping therapeutic empathy expression, and in turn, measurement.

### 4.5. Conclusion

We developed a universal single-item measure of therapeutic empathy both in pictorial and text-based forms. Patient-reported versions demonstrated strong validity, sensitivity to known group differences, and high acceptability. In increasingly time-pressured healthcare environments, this brief, conceptually grounded, and accessible measure offers a pragmatic approach to therapeutic empathy assessment. With further validation, it can support routine therapeutic empathy measurement, strengthen feedback loops in clinical practice and education, and contribute to international efforts to enhance empathic healthcare and improve patient and practitioner outcomes.

## Data Availability

Data cannot be shared publicly because of ethical restrictions, as data contain potentially identifying personal information. Requests for data access should be sent to.

https://drive.google.com/file/d/1DgtYp4JBYAnPucSyt8B7xGMDgCVkENp_/view?usp=sharing

https://drive.google.com/file/d/1BvuhojZwUoI4G1Aj6BIXGNgpLCXA9TIg/view?usp=sharing

## Acknowledgements

The authors would like to acknowledge the participants, without whom the study would not have been possible.

## Supporting information captions

**S1 Appendix. Questions used to screen digital exclusion and illiteracy based on previous research**

**S2 Appendix. Table describing the components of therapeutic empathy.**

**S3 Appendix. Free-text questions used to elicit expert panel feedback for the assessment of content validity.**

**S4 Appendix. Topic guide for semi-structured cognitive interviews.**

**S5 Appendix. Question to assess the construct of clinical neutrality.**

**S6 Appendix. Initial version of the universal single-item scale to measure therapeutic empathy.**

**S7 Appendix. Results of readability tests for initial versions of the scale.**

**S8 Appendix. Participant characteristics: Assessment of content validity.**

**S9 Appendix Participant feedback and modifications made to the scales at each stage of development.**

**S10 Appendix. Participant characteristics: Pre-testing the scale in cognitive interviews.**

**S11 Appendix. Final versions of the universal single-item scales.**

